# Protocol for Readiness of Rehabilitation services integration in Primary Health care: A study in Karnali Province, Nepal

**DOI:** 10.1101/2025.08.19.25334032

**Authors:** Brish Bahadur Shahi, Nista Shrestha, Sunil Pokharel, Thaneshor Paneru, Rita Adhikari, Prakash Upadhayay, Kanchan Thapa

## Abstract

**Introduction:** Globally, 8 million people experience conditions for which rehabilitation services can be beneficial. The services are considered as an essential health service for attainment of universal health coverage. According to the Nepal Health Demographic Survey (NDHS) 2022 report 23 % of the population have some type of functional disability among which 6% people had total functional disability. This study aims to assess the rehabilitation service readiness status, knowledge of health workers, experience of service receivers and policy framework level.

**Methods:** A mixed method study design will be conducted to assess the readiness for rehabilitation services for a period of two starting from July 2024 to 2026. This study protocol clearly aims to examine the status of rehabilitation services, knowledge and skill gap of health workers, experiences of elderly and persons with disabilities for rehabilitation service utilization. Similarly, the perception and experience of working in the health services sector will be assessed through Key Informant Interview (KII). The participants of the current study will be 23 health facilities, >300 health workers working in primary health care in Karnali, convenience sampled population of elderly and disabled population of Karnali. The study team designed a study protocol and conducted pre-testing in a similar setting.

**Ethics and dissemination:** The present study is approved by Nepal Health Research Council(Ref #49, July-21, 2024). Prior to the ethical approval, the study team received approval from the Ministry of Social Development Karnali Province. Informed consent was taken from all the participants involved in this study. All the data collected will be used for study purposes only. The findings of the study will be published as a report from the Publication of the Ministry of Social Development and major findings will be published in Journal.

## Introduction

Globally, 8 million people are experiencing health conditions which require rehabilitation (1). About 2.41 billion people globally could benefit from rehabilitation.(2) Among them, 240 million children are disabled in the world. (3) There has been a rise of cases requiring rehabilitation to 79.4% from 1990 to 202021 and the WHO Western Pacific Region has the highest need of different rehabilitation services. The musculo-skeletal condition with low back pain has the highest prevalence for the need of rehabilitation across the globe. (2)In the South-East Asian Region (SEAR), it has been estimated that approximately 1 in 3 people could benefit from rehabilitation services and 120 million people have been affected by vision loss. There has been a rise of cases requiring rehabilitation to 101% from 1990 to 2021. India has highest number of people that could be benefitted from rehabilitation in SEAR.(2)

Rehabilitation is a set of health interventions that optimize functioning and reduce disability in individuals with health conditions in interaction with their environment.(1) Rehabilitation addresses the diverse range of health conditions, from acute to chronic health conditions, and from physical, mental health, vision and hearing related conditions.(4) Rehabilitation is a highly integrated health service and should be part of the continuum of care of a wide array of health conditions. Rehabilitation aims to achieve and maintain optimal functioning, and functioning can be measured at individual and population level. Moving, seeing, hearing, speaking, communicating and engaging in social interactions are forms of human functioning, and they are crucial for a healthy, dignified, and meaningful life. It needs to be integrated into an array of clinical care, delivered along the continuum. Mostly, it is multi-disciplinary, team-based care, but can be single discipline. Box 1. lists the six categories of interventions for rehabilitation and combinations of these interventions are usually delivered in rehabilitation services.(2)

### Box 1.

**Rehabilitation interventions**

1. Therapeutic techniques and procedures, exercises and training (e.g., manual therapy, range of motion exercises, cognitive behavioral therapy and communication skill training)
2. Physical modalities (e.g., neuromuscular electrical stimulation)
3. Assistive products (e.g., provision and training in the use of wheelchair)
4. Environmental modifications (e.g., installation of ramps, bathroom modifications)
5. Self-management interventions (e.g., education and advice on self-directed training, family and career training)
6. Medicines (e.g., oral non-steroidal anti-inflammatories)

WHO included rehabilitation as an essential health service in the definition of Universal Health Coverage. Rehabilitation is required to prevent further complications, shorten recovery time as well as to improve the physical and mental functioning and wellbeing of individual.(5) The rehabilitation needs of the population, shows variation across countries. It is essential for countries to work on interventions that are proven to improve and maintain functioning of people requiring rehabilitation.

According to WHO, one quarter of the population benefits from rehabilitation services and low back pain being the most prevalent condition requiring rehabilitation in Nepal. Approximately 9.0 million people experienced conditions such as musculoskeletal disorders, neurological disorders, sensory impairments, mental disorders, chronic respiratory diseases and neoplasm that could benefit from rehabilitation.(5) According to Nepal Health Demographic Survey (NDHS)2022 report-23 % of people have some type of functional disability among which 6% people has total functional disability or experienced a lot of functional difficulties.(6) Among children aged 2 -17 years, 10.2% have with a lot functional difficulties or full functional di difficulty.(3) In Nepal it is estimated 8.9% of total population requires assistive products for rehabilitation. Out of those with need of assistive devices, 17.9% are deprived of those services.(7) The census of Nepal 2078 showed 2.25% of total population are living with disability.(8) In Nepal 83% of people with disabilities has no access to rehabilitation services.(9) The census of Nepal showed 3.14% of total population of Karnali are living with disability.(8) The percentage of women and men who have a lot of difficulties or cannot function at all is highest in Karnali Province.(6) The total government investment is only 0.2% out of which only 5% is from government source and 95% is from foreign aid. In the case of rehabilitation services, the service provider to total population ratio is 1:20,000 which is not only low but also unequally distributed.(10) District Health Information Software-2 (DHIS2) is an open source software that facilitates data collection and reporting of information through electronic platforms. Enhancing health information systems is crucial to strengthen rehabilitation services. (11)

Aligned to the Public Health Service Act 2018(12) and National Health Policy 2019(13), Karnali Province Health Policy 2019(14) has identified rehabilitation as the health service. The provincial government of Karnali has established the physiotherapy service in all district hospitals. Likewise, Province hospital Surkhet is being developed as the referral hospital for rehabilitation, as evident by the fiscal year 2079/80 budget allocations for the establishment of Prosthetic & Orthotic Service. (15) Even though the province has made encouraging progress, rehabilitation is still absent from primary healthcare, indicating a significant unmet need for the program. In this context, the province recently launched its rehabilitation strategy 2024-2030 in which integration of rehabilitation into primary health care is a priority. In a pursuit of realizing rehabilitation at primary health care, the health division of MoSD intends to assess the rehabilitation service readiness status, knowledge of health workers, experience of service receivers and policy framework level. The system covers all the six building blocks of health applied to primary health care context.

## Material and methods

### Study Setting

Karnali province being the remote province in Nepal comprises 79 local level governments and most of the areas are composed of hills and mountains(16). People still struggle with the use of primary health care services, and the state of rehabilitation services has not yet been thoroughly investigated.

The first part of the readiness assessment was conducted in health facilities of Karnali province (Table. 1). During this stage, a self-administered questionnaire was used to interview the rehabilitation as well as health service providers of these medical facilities. People with disabilities and the older people will be interviewed in the second section. Key stakeholders, including public organizations, INGOs, NGOs, and business partners like Handicap International (HI), National Federation of Disabled-Nepal(NFDN) Federation, etc., that are involved in rehabilitation in Karnali Province will be interviewed for the third section of the study using a qualitative approach.

**Table 1.** Distribution of health facilities by district of Karnali Province, Nepal.

| S.N | District | Community Hospital | Basic Hospital | Primary Health Care Centre | District Hospital | Remarks |
| --- | --- | --- | --- | --- | --- | --- |
| 1 | Surkhet | 1 | 0 | 3 | 0 | Karnali Province Hospital |
| 2 | Salyan | 0 | 2 | 2 | 1 |  |
| 3 | Dailekh | 0 | 4 | 2 | 1 |  |
| 4 | Jagarkot | 2 | 1 | 1 | 1 |  |
| 5 | Rukum West | 1 | 4 | 1 | 1 |  |
| 6 | Kalikot | 1 | 5 | 0 | 1 |  |
| 7 | Jumla | 0 | 0 | 1 | 0 | Karnali Academy of Health Sciences |
| 8 | Mugu | 1 | 0 | 1 | 1 |  |
| 9 | Dolpa | 0 | 0 | 0 | 1 |  |
| 10 | Humla | 1 | 0 | 0 | 1 |  |
| **Total** |  | **7** | **16** | **11** | **8** | **2** |

The health service utilization statistics for Karnali Province in the fiscal years 2022/23 and 2023/24 *(Supplementary Table 1)* shows a general trend in the number of service users across most age groups, with notable decreases in individuals aged 18-59, >59, and 6-17 years. The service utilization trend of people with non-communicable diseases, accidents/injuries, and violence-related cases also declined, while cases categorized as “Others” and those marked “Don’t Know” increased. There was a significant rise in the need for mobility aids, with more patients receiving these devices compared to the previous year. The number of assistive products provided surged, particularly those sourced from hospitals and self-purchased. Those with the diseases of the musculoskeletal system remained the largest consumer group of rehabilitation, though slightly reduced, while consumers with respiratory and genitourinary diseases increased. Rehabilitation services, especially physiotherapy, remained consistent, but the provision of assistive devices experienced a substantial increase. Physical disabilities remained the most common, but there was a notable rise in mental/psycho-social disabilities.

### Study Design

A mixed study parallel convergent design will be adopted for this study. Consolidated criteria for reporting qualitative studies (COREQ): 32-item checklist will be used for data assessment for qualitative findings.

### Sampling

A census of all 23 basic hospitals will be conducted as part of this study. This study will include all hospitals that provide basic primary health care services and are operated at the local level in the province. Health workers were selected conveniently based on their interest to respond to the questionnaire sent to them via link in their email. Similarly, the aging elderly and people with disabilities living in the province will be selected conveniently. For qualitative study, the required number of participants will be selected purposely. The attached STROBE design shows the process of selection of the participation in this study.

**Figure 1.**
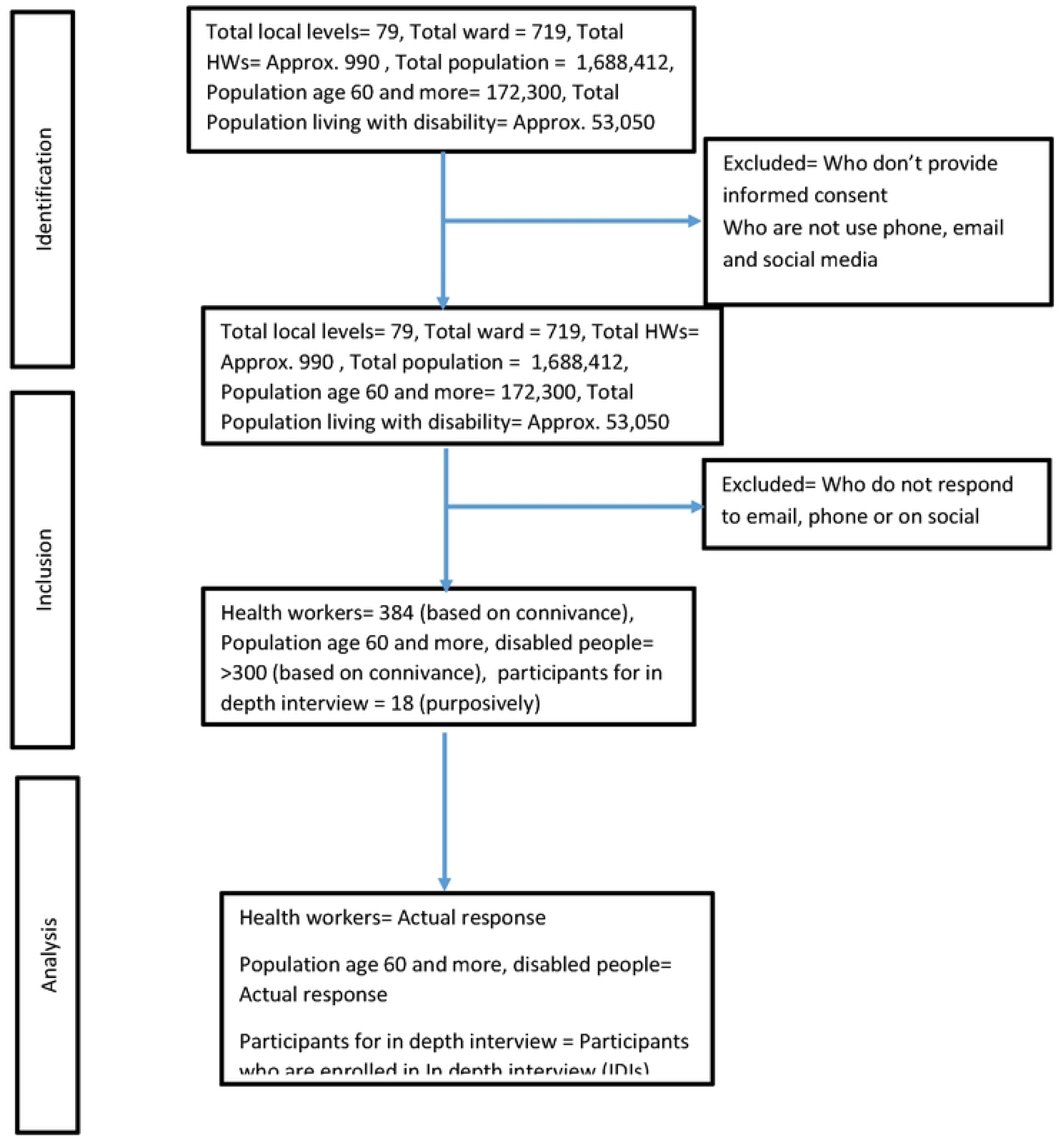
STROBE flow chart (STROBE: Strengthening the Reporting of Observational Studies in Epidemiology).

### Study Participant

This study will be carried out among the health service provider or manager working in a hospital of Karnali province, Nepal. The service provider who has at least a year experience and working in the health facility continuously will be recruited upon informed written consent. Thus, for the readiness assessment part, 23 local level primary hospital managers will be the study participants. Similarly, health workers working in health service delivery points of the province will be the study participants for knowledge related information. Furthermore, older people and persons with disability working in the province will be assessed to examine their lived experience of the service.. Moreover, officers working in health service directorate, health coordinator of local levels, health service province, female community health volunteer, officer of social development ministry, health service manager of the district level, local level representative, development partner representative will be interviewed for key informant interview. At present, after the implementation of constitution of Nepal 2015, the health system of the country is undergoing several changes which also provided various opportunities and challenges during the reform. (17) Similarly, health workers who wish to provide information and feedback can write their response in the written designed questionnaire at the place of their convenience.

### Study tool

A standardized pre-validated tool will be used for this study. A zero draft of the tool was developed based on the WHO SARA and Nepal Health Facility Tool for the other health services. Similar rehabilitation technical guidelines, DHIS2 system, available training at the national level were referred to derive the domains and tracer areas. Review of available documents were done for this purpose. Based on the review, pre-testing, expert consultation and consultation with the key stakeholder in the province. The readiness checklist (*Supplementary file 2*) was developed which will be used in this study after considering the pre-testing in the selected municipal hospitals of the province. The key indicator for the tool will be as in table. 2.The study tool will cover mainly 10 domains.

**Table 2.** Distribution of Key indicators in readiness assessment questionnaire.

| Domains | Tracer areas |
| --- | --- |
| Scope of work | Rehabilitation scope defined for municipal hospital<br>Screening pathways for health conditions established with other public health programs such as NCDs, NTDs, senior citizen program, nutrition, ECD , disability, MNCH Health profile of the municipality covered rehabilitation |
| Human Resources (Core) | Rehabilitation staff availability<br>Health worker trained in basic rehabilitation<br>Health worker trained in Training on Assistive Product(TAP) |
| Quality | Minimum service standard for rehabilitation at municipal hospital<br>Rehabilitation Quality assurance guideline/protocols for municipal hospital |
| Service utilization status | Number of cases attending municipal hospitals for any kind of health services: Low back pain, vision loss, hearing loss, COPD, fractures, Neck pain, osteoarthritis, other musculoskeletal injuries , Stroke, Traumatic brain injury Amputation, Developmental delays, cerebral palsy, autism , spinal cord injury , pelvic organ prolapse |
| Therapeutic equipment | Exercises mattress, Theraband , dumbbells, sandbags, hydro collated packs, cold packs , balance boards, shoulder wheel, Hand exerciser , ankle exerciser , age appropriate toys, posture correction mirror , gym balls, putty clay, couches, pillows, wedge, foam roll and incentive spirometer |
| Assistive products | Transfer board, grab bar, portable ramp, crutches, walkers, walking sticks, therapeutic foot wear, spectacles, magnifier, pressure relief mattress, catheters, urine bag, bed pan, pressure relief mattress/cushion, wheelchair and toilet chair |
| Screening equipment | Pen torch, tape measure, goniometer, weighing scale, tuning fork, knee hammer, height measuring equipment, stethoscope, BP set , Pulse oximeter |
| Infrastructure (Core) | Secure space designated for the delivery of the service,<br>Registration, toilet and rehabilitation room accessible |
| Service reporting & recording | HMIS/DHIS2/ rehabilitation module available for primary health<br>Facility enrolment in HMIS/DHIS2 rehabilitation module<br>Staff trained in HMIS/DHIS2 rehabilitation module |
| Financing | Rehabilitation funded in basic health care package<br>Municipal willingness to cover rehabilitation through its own fund in municipal hospital |
|  | MoSD willingness to top-up the funding of rehabilitation in municipal hospital |

**Table 3.** Distribution of findings of province level consultation on rehabilitation services in province.

| S.<br>N | Issues | Recommendation |
| --- | --- | --- |
| 1 | Lack of dedicated skilled manpower for rehabilitation services | Recruitment of permanent physiotherapy personnel, Diploma courses to local students in physiotherapy, need based ONM, Province need to take lead role for recruitment process |
| 2 | Problem in service delivery due to lack of awareness of people | More awareness programs to be conducted to increase programme coverage, Timely production and dissemination of IEC materials regarding rehabilitation services |
| 3 | Low budget on rehabilitation services | Advocacy, allocation and implementation of evidence based budgeting |
| 4 | Lack of adequate space for physiotherapy and rehabilitation services | Establishment of separate and dedicated space for rehabilitation services |
| 5 | Lack of alternative staff for smooth running of rehabilitation services | At least needed two dedicated staff at each service point |
| 6 | Lack of orientation and training to staff | Timely and need based training need to be conducted periodically, Other health staff like HA, Staff nurse need also to be trained for continuous delivery of rehabilitation services |
| 7 | Problem in referral system | Strengthening referral system between different health institutions to rehabilitation unit |
| 8 | Lack of availability of adequate rehabilitation equipment's | Logistic supply as per pull system, increasing funds for procurement of need based rehabilitation materials, taking supports of EDPs to fulfill need of materials |
| 9 | Lack of ownership of services by community, health section and other stakeholders | Advocacy and timely orientation to all concerned stakeholders |
| 10 | Inefficient implementation of formulated guidelines | Proper research while developing policies and management protocols |
| 11 | Geographical barriers | Transfer of basic rehabilitation skills to health post, CHU, BHSC health personnel's to offer basic services |
| 12 | Understock of assistive devices | Procurement and Stock piling of needed devices |
| 13 | Poor coordination between concerned authorities | Development of efficient coordination, communication mechanisms among all concerned authorities |
| 14 | Lack of proper incentives to service providers | Provisions to make necessary increase in incentive and allowances |

### Health Workers Perspectives

The health workers questionnaire (*Supplementary file 3)* will assess the socio-demographic background, knowledge, their current practices for referral, felt need based on their clinical practice will explore. A total of 27 questions are included in this questionnaire. As this is a self-administered questionnaire filled in Google form, health workers are free to answer as per their wish.

### Persons with Disability and Older people Perspectives

This study will consider persons with disabilities, aging and elderly perspectives to be included in the study. As a major service user their view points are considered important to consider in this study. Therefore, a questionnaire is developed and will be sent to the elderly and people with disabilities. The data collection will be done through the Google form, however due to some inclusive requirements minor assistance during the data collection will be provided. The nature of such assistance will be recorded however the study team/ volunteer will ensure that there will not be any bias during the data collection. The questionnaire consists of 35 questions covering socio-demographic, their knowledge, user history, referral challenges faced, health workers behaviors etc (*Supplementary file 4)*.

### Key Informed Perspectives

The key informant for this study will be identified which includes province level health care manager, district level health care manager, clinician, local level health coordinator, health workers working in service delivery point, service user, female community health volunteer, local level representatives, and other key stakeholder identified through the study period. All the information will be collected through the KII guideline developed in Nepali and English (*Supplementary file 5*). All the data collection will be done up to the saturation level.

### Findings from Province Levels Consultation

As a part of validation and further needs of this kind of the study, province level consultation was organized on 16-17^th^ May 2024 in the Karnali province. The workshop further explored the current situation of the rehabilitation services in the Karnali province. Based on the findings of the province level workshop, further modification of the tool will be done (*Supplementary file 6*). The findings of the province level workshop are presented in the table.

### Study tool pretesting

The study tool will be pretested in two health facilities of the province which will be selected purposely. The basic hospital of the Surkhet and Dailekh will be selected purposely. Based on the findings of the pre-testing, further modification of the tool will be done. Similarly, the rest of the tools and questionnaire (*Supplementary 3-5*) will be pre-tested in Surkhet district. Based on the findings of the pre-testing the tool will be further modified. Further, language and technical modification will be done based on expert suggestion and advice closely adhering to pre-testing findings.

### Data collection in the field

The data collection in the present study will be done through the online platform Google form. The form will be developed in the local Nepali language. A link to the study participants will be sent through an email. The study participants (health service provider/manager) of the selected health facility will provide the response in the provided Google sheet.

### Data processing / entry

The data will be collected through the validated Google form which will be filled up by the study participants. As the study participant directly enters the provided information in the Google sheet, no further entries are required. Furthermore, a strong validation option is kept in the Google form for the data capturing. On a daily basis, the summary of the field progress will be monitored and shared to the study team.

### Data analysis

The data analysis will be done as per the domain and tracer indicator selected. The inferential and descriptive analysis will be done in the R studio. The analytical statistics will be reported in 95% confidence along with their frequency and percentage. The findings of the study will be presented in tables and figures.

### Ethical consideration

The present study received an ethical approval from Nepal Health Research Council (Ref #49, July-21, 2024). Prior to the ethical approval, the study team received an approval from the Ministry of Social Development Karnali Province. In this study, the written informed consent will be obtained from the health facility manager/service province. The informed consent form in local Nepali language will be sent to the health facility in email along with the link for the data collection. The informed consent form will clearly state about the purpose of the study, harms-benefits of the study, voluntary participation, etc. The health facility manager will sign the form and return to the research team with a signature. All the data collection in this study will be done in local Nepali language however the simultaneous translated version is available for the validity and proper understanding of the study participants.

### Strengths and limitation

This is the first of this kind of the study initiated by the provincial government in Karnali, Nepal. The research team is composed of public health professionals, epidemiologist, health administrator, policy and planning experts, and physiotherapists. All the basic hospitals in the province who are providing primary health care service will be included in the study. We used the pre-tested validated questionnaire tool (*Supplementary file 2-7*) in this study. Prior to the finalization of the tool, we consulted with the stakeholder in the province for the situation analysis. Based on the situation analysis, the further modification of the study protocol was done. Furthermore, the tool was reviewed by different experts of the primary health care, physiotherapy and public health services from the government and non-government sector. In this study, we proposed a pre-tested, validated tool for data collection which can be further replicated in other settings where the basic rehabilitation services were planned to be operationalized through the primary health care delivery points. In order to validate the tool, we will consider feasibility, internal consistency and sensitivity of the tool. Based on the pre testing, modification will be done on the questionnaire. At the national and global scale, integration of rehabilitation at primary health care is a key priority. Therefore, this tool is expected to add a strong resource in an ongoing global effort to bring rehabilitation to primary health care.

### Study timeline

The total period of the present study will be one year starting from May 2024 to April 2025. All the activities have been planned to track the progress of the study and smooth project operation (*Supplementary file 7*). The study will share its period report to Nepal Health Research Council (NHRC) and Ministry of Social Development (MoSD) Karnali Province. Upon the completion of the report, the final report will be handed to MoSD and NHRC.

## Discussion

This study will be conducted with the objective to determine the challenges, gaps and opportunities of integrating rehabilitation services in primary health care services in Karnali Province. The study aims to ascertain the situation of health care facilities of basic hospitals for providing rehabilitation services and explore the role of health care provider, patients and administrator role in integrating rehabilitation services in primary health care services in Karnali province, Nepal. Along with it, the study is designed to determine the needs and perspective of the person requiring special needs (disabled person/aging) and their current knowledge on rehabilitation among primary health care regarding rehabilitation services needs and referral system in Karnali province. With the vision of “A quality of health service within reach of all residents of Karnali: happy, alert and conscious Karnali”, Health Sector Rehabilitation Action Plan 2080-2087 has been formed in Karnali Province(15). Our study is outlined in accordance to support the goal envisioned by Karnali Health Policy 2078(14), Health Sector Strategic Action Plan(15), and Rehabilitation Strategic Action Plan, 2080-2087(18).

Consideration of both the qualitative components and qualitative factors, along with a policy and program implementation and evaluation makes our study a comprehensive study for integration of rehabilitation services in primary health care. Furthermore, our qualitative research design allows flexibility during the iterative process of consultations with key stakeholders and experts. Importantly, collaborating with key stakeholders during the development process will help in the uptake and ownership of the results disseminated by the research process.

The pre-testing of the questionnaire will be done based on the selected sites. The rehabilitation facilities, primary health care center, provincial hospital, district public health office and other external development partners which can be reviewed in provincial annual report(16) working for rehabilitation in Surkhet district. The survey form developed from Google survey will be asked to be filled and the validity and reliability of the questionnaire will be checked. The KII questions will also be checked through the interview process. The DHIS II data of FY 2023/24 of Karnali province displayed the need for rehabilitation service to be highest (63.2%) in the age group of 18-59 years followed by >59 years (21.9%). The musculo-skeletal and connective tissue related disability was found to be maximum (70.5%) followed by injuries and external causes (9.7%). The mobility aid had the highest need (98.4%) among other assistive devices. The physiotherapy service had maximum coverage (86.4%) among all other services. Among all the types of disability the physical disability was most prevalent (97.31%) in Karnali province (*Supplementary Table 1*).

The present study is the first study of this type initiated by the provincial government of Karnali, Nepal. It involves the qualitative and quantitative methods using the mixed methods approach. The health workers, aging, elderly and disabled people living in the province are open to provide their feedback. In order to record, we will include an option for open ended questions in each form (supplementary file). Similarly, the quantitative data will be analyzed through the deductive approach and validated with quantitative findings. Though there are various frameworks, guidelines and protocols for assessing health system readiness(19), WHO guideline for quality assessment for pharmaceuticals(20), readiness assessment however there is limited protocol to assess the rehabilitation readiness. A paper published assesses the number of rehabilitation centers in SEAR region(21) however, the country and province specific information is limited on it. Such health service information is crucial for other health conditions such as stroke management.(21) Furthermore, there are various assessment reports for assessing health service readiness for maternal and child health(22), health system gaps for cardiovascular diseases(23), sustainable development baseline report in Karnali(24), and others in the provinces. Being a rural province of Nepal, the emphasis on primary health care and its integration with rehabilitation is important.

However, despite these strengths, this study will have some limitations. The study will be conducted among the participants who provide responses which will be shared to them. If someone with limited literacy over digital media will be automatically excluded from the analysis. Similarly, the qualitative data will be collected purposely and will be mainly concerned in the highly accessible places of Surkhet. Therefore, the information of remotely inaccessible areas will be automatically excluded.

In summary, this protocol will capture the challenges, opportunities, knowledge gap, skills gaps, equipment status and rehabilitation service readiness in Karnali province through a parallel convergent mixed methods design. The findings will have policy level implications to integrate rehabilitation services in the province.

## Data Availability

The data of the previous year service statistics is reported in supplementary file 1.

NA

## Supporting Information Supplementary File

***Supplementary Table 1***. Distribution of service statistics of the province by service users, providers, services provided, diagnosis and needs of Karnali Province in Fiscal year 2022/23-2023-24.

***Supplementary File 2***. *Readiness Assessment Questionnaire (English and Nepali)*

***Supplementary File 3***. *Questionnaire for current knowledge on rehabilitation among primary health care regarding rehabilitation services needs and referral system in Karnali province, Nepal (English and Nepali)*

***Supplementary file 4***. *Questionnaire on Challenges and Utilization of Rehabilitation Services Among the Elderly and Differently abled person (English and Nepali)*

***Supplementary file 5***. *Key Informant Interview Guideline for different levels (English and Nepali)*

***Supplementary file 6***. *Guideline for Province Level Consultation for Rehabilitation Service Status, Challenges and Opportunities (English and Nepali)*

***Supplementary file 7***. *Timeline of the present study*

## Acknowledgement

The study team would like to thank Nepal Health Research Council (NHRC) for timely approval of our research protocol. We would like to thank all the study participants willing to take part in this study. We would like to thank the Ministry of Social Development (MoSD) and Province Health Service Directorate Surkhet for their continuous motivation and support to conduct this study.

## Contributions

BBS and KT conceptualized the study, wrote the first protocol. NS, SP,BNA,TPP, RA, PU reviewed the protocol, reviewed the literature, collected the baseline information, and analyzed the data. BBS and KT finalized the tools, protocol and obtained ethical approval. All the authors approved the first draft of the protocol.

## Supplementary file

The present study was completed within 1.5 year. The detail is presented in the following Gantt Chart. Table. Timeline for Proposed Research (June 2024-Dec 2025)

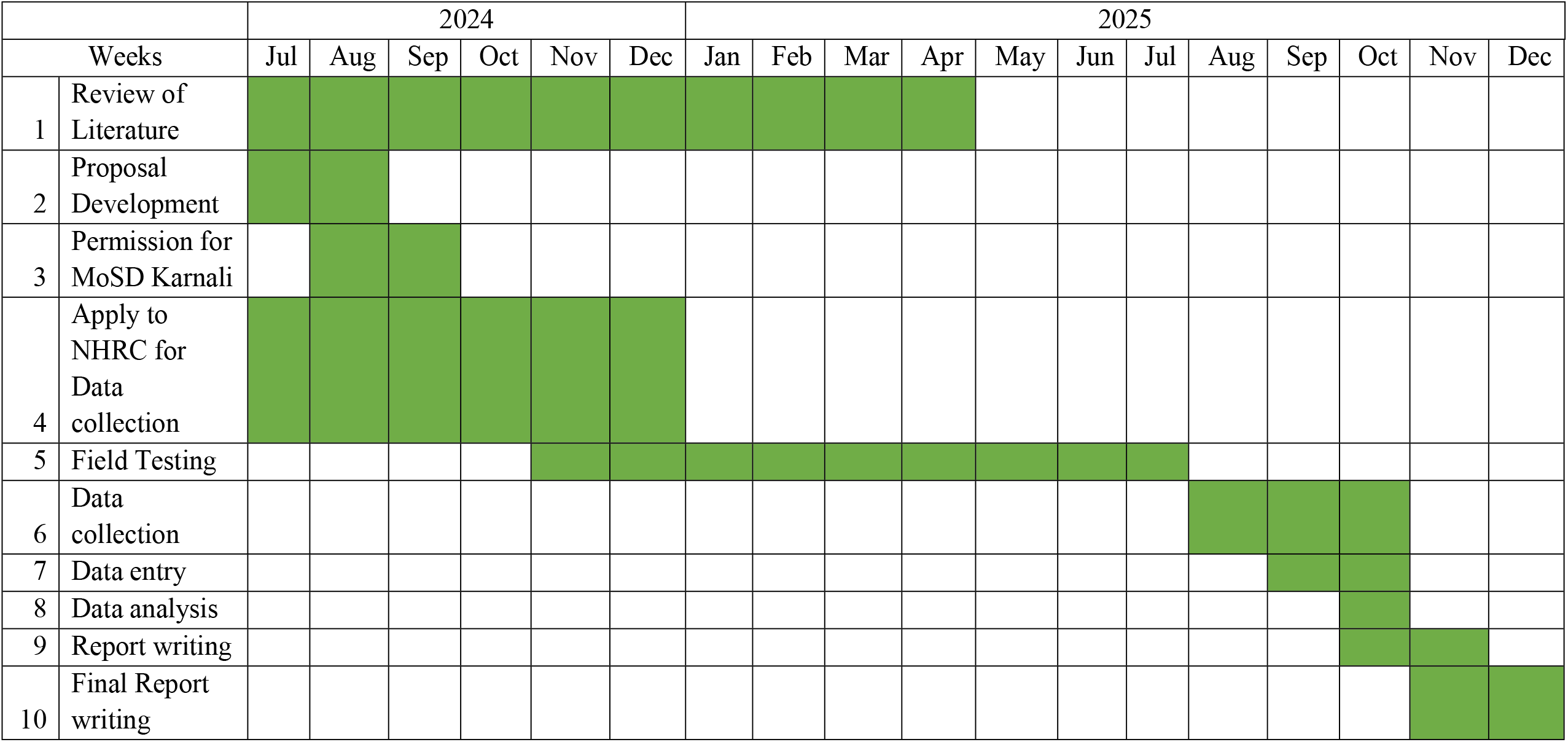

